# Modeling In-Hospital Mortality Among Patients Undergoing Percutaneous Coronary Intervention with Acute Myocardial Infarction Complicated by Cardiogenic Shock Receiving Mechanical Circulatory Support

**DOI:** 10.1101/2025.11.24.25340896

**Authors:** Nathan C. Hurley, Bobak J. Mortazavi, Sanket S. Dhruva, Joseph S. Ross, Che G. Ngufor, Jeptha P Curtis, Harlan M. Krumholz, Nihar R. Desai

## Abstract

Acute myocardial infarction complicated by cardiogenic shock (AMI-CS) is a heterogeneous clinical syndrome associated with substantial morbidity and mortality. We developed a machine learning-based mortality model to identify features and patient subgroups associated with the largest change in mortality risk when evaluating treatment with Impella devices or intra-aortic Balloon Pump (IABP). Our cohort comprised 369 sites and 15,796 patient visits to the cardiac catheterization laboratory from the National Cardiovascular Data Registry. The estimated population mean excess mortality effect of treatment with Impella devices vs IABP was 10.4 ± 0.8%. However, we identified clinical subgroups of 282 patients for whom a decreased risk of mortality was associated with use of Impella as compared with IABP. Those patients were on average younger, presented with higher systolic blood pressure, higher rate of salvage percutaneous coronary intervention, higher initial creatinine, and lower hemoglobin. While Impella devices were associated with higher mortality risk overall, certain clinical profiles were associated with lower risk, illustrating heterogeneity of treatment effects.

## Introduction

Acute myocardial infarction (AMI) complicated by cardiogenic shock (AMI-CS) is associated with substantial morbidity and mortality. However, clinical outcomes among patients with AMI complicated by cardiogenic shock (AMI-CS) have not substantially improved over time^1–3^, with estimated mortality as high as 60%^4^. AMI-CS is a heterogeneous clinical syndrome, encompassing heterogeneous patients with differing degrees of clinical presentation and symptom or condition severity, which may potentially be responsive to differing interventions and strategies^5, 6^. However, many prior studies may have failed to adequately account for this clinical heterogeneity. The absence of a clinical benefit of a particular strategy or intervention may have been related to suboptimal risk adjustment. Correctly identifying these subgroups may therefore allow for identification of interventions leading to benefit in those subgroups.

One intervention used for these patients is mechanical circulatory support (MCS) devices. Approximately 40% of patients with AMI-CS are treated with MCS devices^7^, most commonly intra-aortic balloon pumps (IABPs) or Impella devices, which are intravascular microaxial left ventricular assist devices (LVADs)^7^. However, clinical trials comparing IABP and Impella devices with each other have been small, underpowered, and provided inconsistent results^8, 9^. Multiple large observational studies found Impella devices were might be associated with significantly increased mortality risk compared to IABP, and these studies have had disagreement upon risks and benefits ^1, 10–12^. More recently, however, the DanGer Shock trial found consistent improvement with microaxial flow pumps. Investigators found that microaxial flow pumps lowered risk of death in STEMI-related cardiogenic shock over standard care, even with adverse events occurring more frequently in this same group of patients ^13^. The investigators of that trial found that their patient population was more homogeneous, likely leading to the identified success of the microaxial flow pump in reducing risk of death. This suggests that patient heterogeneity has been a leading factor towards inconsistent results.

An improved understanding of heterogeneity of patients and associated risk and benefit may be a critical factor to determine the patient subgroups for whom various MCS devices may be associated with improved clinical outcomes among patients with AMI-CS. Understanding this heterogeneity and accounting for it in modeling may help develop hypotheses for prospective randomized evaluation of MCS devices in clinical scenarios where they may be more likely to demonstrate reduced mortality risk. We hypothesized that adjusting for patient characteristics would find the homogeneous subgroups of patients for which risk adjustment models would provide more tailored decision-making about MCS device selection. Accordingly, we developed a machine learning-based mortality model to identify features and patient subgroups that were associated with the largest change in risk when evaluating treatment with specific MCS devices.

## Results

### Cohort, Model, and Covariates

Our cohort comprised 369 sites and 15,796 patient visits to the cardiac catheterization laboratory from the American College of Cardiology’s National Cardiovascular Data Registry CathPCI and Chest Pain-MI registries, as previously described^1, 7^. In-hospital mortality risk was estimated using machine learning. Specifically, we employed a stratified five-fold cross-validation procedure to train the extreme gradient boosting machine (XGBoost)^14^ algorithm to predict in-hospital mortality. We used this model because of how the various decision trees trained lead to a divide-and-conquer strategy of patient populations, modeling in each subgroup with multiple interaction effect terms, which has been shown to improve risk prediction in registry data for other models^15–17^. Model hyperparameter (learning rate, maximum depth of each learner, and number of learners) tuning was conducted through grid search on the training set within the cross-validation. For the final models, the hyperparameter tuning lead to using a learning rate of 0.3, maximum depth of each tree estimator of 6, and with 100 trees. The initial set of candidate features was provided in a prior study and described there ^1, 7^. After feature selection, the base model included patient age, sex, percutaneous coronary intervention (PCI) status (elective, urgent, emergent, salvage), recent history of cardiac arrest, initial creatinine, initial hemoglobin, stenosis present in left main coronary artery (percent), stenosis present in proximal left anterior descending coronary artery (percent), presence of multivessel disease, systolic blood pressure at first medical contact, recent history of thrombolytic therapy, and findings on first electrocardiogram (ECG) (no ST elevation, ST elevation, new left bundle branch block, or isolated posterior MI). The cohort characteristics are noted in **Table 1** and their association with mortality and major bleeding in **Table 2**. This study was considered exempt by the Human Research Protection Program at Yale University under IRB #0607001639.

**Table 1.** Patient Characteristics. All fields are count (n) and percentage, unless otherwise noted as mean and standard deviation.

|  | **All Patients**<br>**(n=15,796)** |
| --- | --- |
| **Age (years)** | 65.7 (± 12.6) |
| **Male Sex** | 10,618 (67.2%) |
| **SBP at First Medical**<br>**Contact (mmHg)** | 114.8 (± 48.2) |
| **Any Cardiac Arrest** | 3,731 (23.6%) |
| **PCI Status** |  |
| Elective | 102 (0.6%) |
| Urgent | 2,265 (14.3%) |
| Emergent | 12,170 (77.0%) |
| Salvage | 1,259 (8.0%) |
| **Initial Creatinine**<br>**(mg/dL)** | 1.44 (± 1.28) |
| **Left Main Coronary**<br>**Artery Stenosis (%)** | 12.2% (± 25.1%) |
| **Multivessel Disease** | 8,751 (55.4%) |
| **Initial Hemoglobin**<br>**(g/dL)** | 13.7 (± 2.2) |
| **Proximal LAD Stenosis**<br>**(%)** | 46.5% (± 43.5%) |
| **Findings on Initial ECG** |  |
| No STEMI | 3,210 (20.3%) |
| STEMI | 12,258 (77.6%) |
| LBBB | 181 (1.1%) |
| Posterior MI | 147 (0.9%) |
| **Thrombolytic Therapy** | 556 (3.5%) |
| **MCS Device Utilization** |  |
| IABP Only | 4,919 (31.1%) |
| Impella devices Only | 1,686 (10.7%) |
| Medical Therapy Only | 9,191 (58.2%) |
| **Major Bleeding** | 2,212.0 (14.0%) |
| **Mortality** | 4,042 (25.6%) |

**Table 2.**
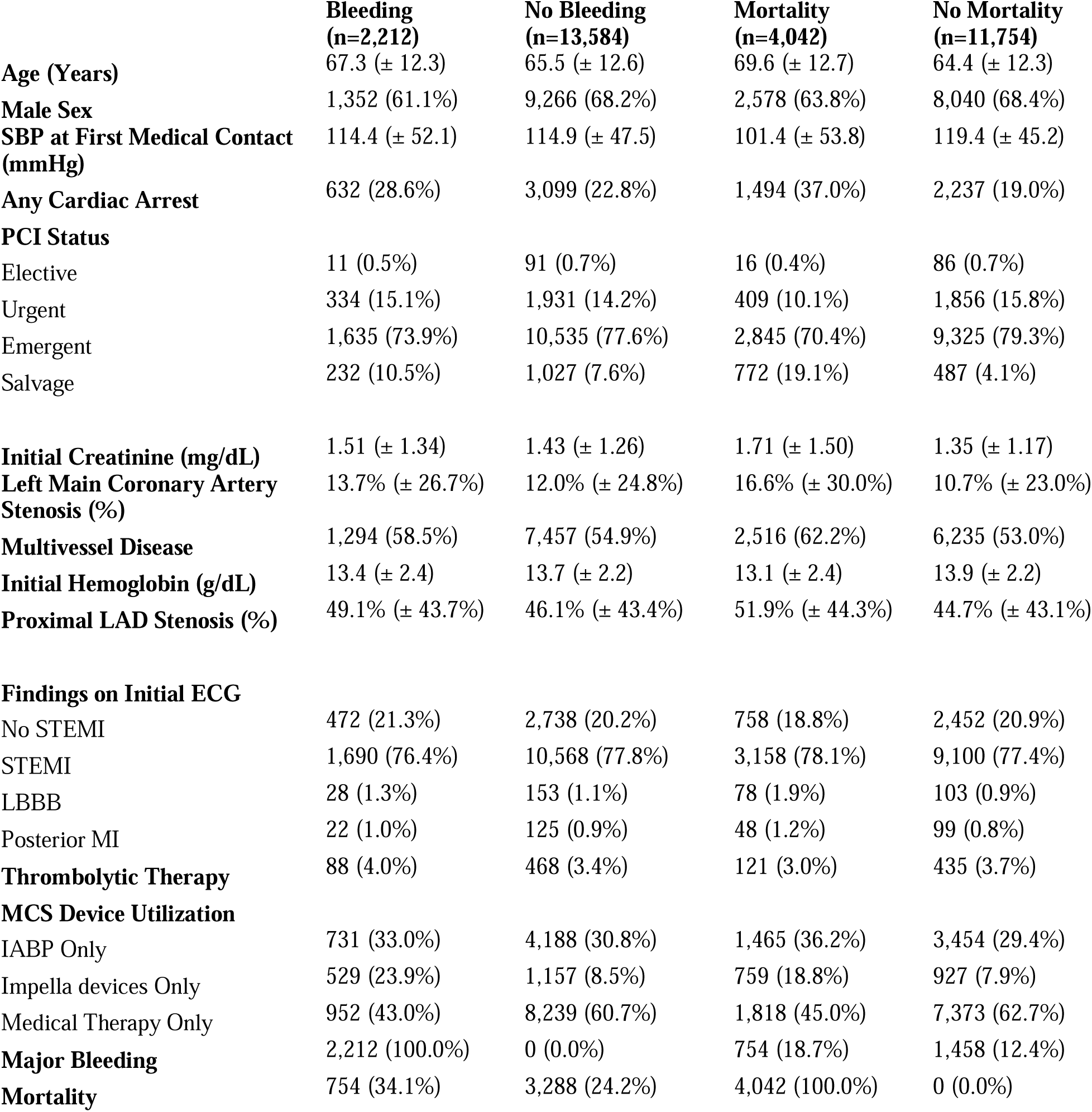
– Raw Association of Patient Characteristics with In-Hospital Major Bleeding and Mortality.

### Model Performance

We measured model performance by the area under the receiver operating characteristic curve (AUROC), for evaluating model discrimination, the average precision score, which calculates the area under the precision recall curve for evaluating model positive predictive performance, and the model Brier Score, for evaluating model calibration ^18, 19^. The base model predicted mortality with a mean area under the receiver operating characteristic curve (AUROC) of 0.747 ± 0.002 (standard deviation) (**Table 3**) and the average precision score of 0.517 ± 0.04. Adding the MCS device utilization variable improved performance to 0.754 ± 0.025 AUROC and 0.519 ± 0.047 average precision score. Adding the major bleeding outcome as a risk factor for mortality resulted in an AUROC of 0.748 ± 0.022 and an average precision score of 0.511 ± 0.043. Adding both bleeding and MCS device utilization achieved a performance of 0.761 ± 0.022 AUROC and an average precision score of 0.522 ± 0.037.

**Table 3.** Model AUROCs.

|  | **AUROC** | **AUPRC** | **Brier Score** |
| --- | --- | --- | --- |
| Base Model | 0.747 ± 0.024 | 0.517 ± 0.043 | -0.164 ± 0.009 |
| Base Model + MCS Utilization | 0.754 ± 0.025 | 0.519 ± 0.047 | -0.164 ± 0.010 |
| Base Model + In-Hospital Major Bleeding | 0.748 ± 0.022 | 0.511 ± 0.043 | -0.165 ± 0.008 |
| Base Model + MCS Utilization + In-Hospital Major Bleeding | 0.757 ± 0.022 | 0.522 ± 0.037 | -0.163 ± 0.009 |

### Estimated Mortality Risk Difference with MCS Utilization

Using the MCS device utilization model, we calculated the risk differences for Impella devices vs IABP (**Figure 2 and Table 4)**. This figure identifies the population-level estimated risk difference in mortality for patients treated with Impella devices vs IABP. The estimated population mean excess mortality effect of treatment with Impella devices vs IABP was 10.4 ± 0.8%,

**Table 4.**
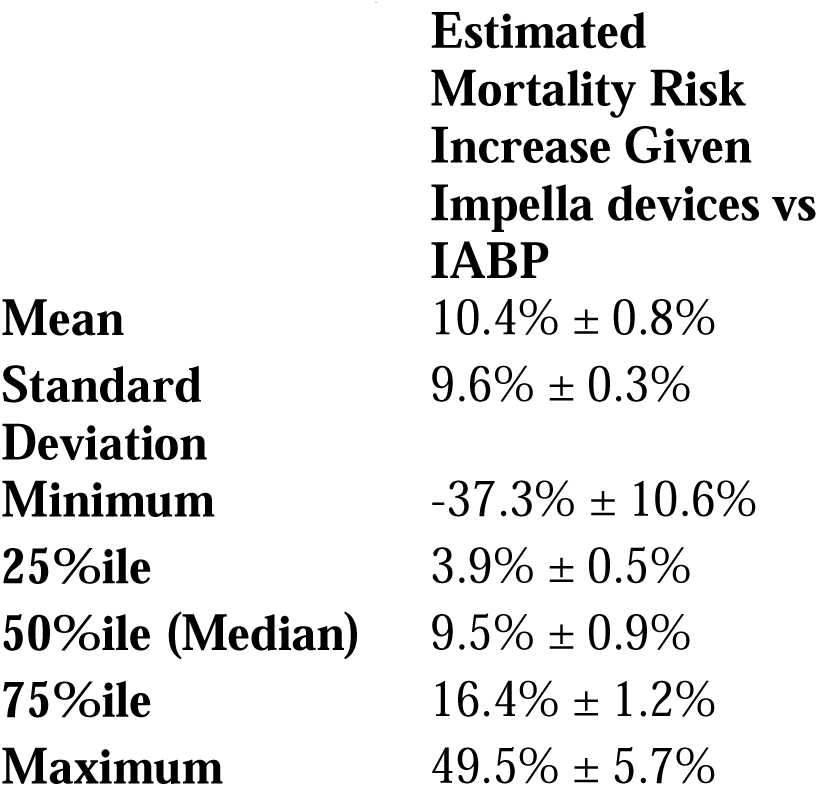
Summary statistics of estimated risk differences.

|  | **Estimated Mortality Risk Increase Given Impella devices vs IABP** |
| --- | --- |
| **Mean** | 10.4% ± 0.8% |
| **Standard Deviation** | 9.6% ± 0.3% |
| **Minimum** | -37.3% ± 10.6% |
| **25%ile** | 3.9% ± 0.5% |
| **50%ile (Median)** | 9.5% ± 0.9% |
| **75%ile** | 16.4% ± 1.2% |
| **Maximum** | 49.5% ± 5.7% |

Taking a single test fold^1^, there were 282 (7.2%) patients who had an estimated decreased risk of mortality when hypothetically managed with Impella devices compared to IABP (**Table 5)**. **Table 5** also presents data from a cohort of equal size from the opposite tail: the 282 patients with the largest increase in mortality risk given Impella devices vs IABP, to identify cases associated with lower risk based upon MCS device choice.

**Table 5.** Descriptive statistics of populations with largest risk shifts given Impella devices vs IABP.

|  | **Most likely to be associated with in-hospital mortality benefit from Impella devices compared to IABP (n=282)** | **Most likely to be associated with in-hospital mortality benefit from IABP compared to Impella devices (n=282)** | **P Value** |
| --- | --- | --- | --- |
| **Age (Years)** | 66.6 (± 14.9) | 70.7 (± 10.0) | < 0.001 |
| **Male Sex** | 174 (61.7%) | 171 (60.6%) | 0.86 |
| **SBP at First Medical Contact (mmHg)** | 130.6 (± 58.4) | 102.0 (± 52.5) | < 0.001 |
| **Any Cardiac Arrest** | 81 (28.7%) | 67 (23.8%) | 0.21 |
| **PCI Status** |  |  |  |
| Elective | 9 (3.2%) | 0 (0.0%) | < 0.001 |
| Urgent | 128 (45.4%) | 19 (6.7%) |  |
| Emergent | 110 (39.0%) | 246 (87.2%) |  |
| Salvage | 35 (12.4%) | 17 (6.0%) |  |
| **Initial Creatinine (mg/dL)** | 2.41 (± 2.59) | 1.25 (± 0.69) | < 0.001 |
| **Left Main Coronary Artery Stenosis (%)** | 15.4% (± 28.8%) | 13.1% (± 26.1%) | 0.26 |
| **Multivessel Disease** | 163 (57.8%) | 157 (55.7%) | 0.67 |
| **Initial Hemoglobin (g/dL)** | 12.9 (± 2.2) | 13.7 (± 2.3) | < 0.001 |
| **Proximal LAD Stenosis (%)** | 52.1% (± 39.6%) | 26.3% (± 40.8%) | < 0.001 |
| **Findings on Initial ECG** |  |  |  |
| No STEMI | 137 (48.6%) | 37 (13.1%) | < 0.001 |
| STEMI | 141 (50.0%) | 240 (85.1%) |  |
| LBBB | 1 (0.4%) | 3 (1.1%) |  |
| Posterior MI | 3 (1.1%) | 2 (0.7%) |  |
| **Thrombolytic Therapy** | 6 (2.1%) | 6 (2.1%) | 1 |
| **MCS Device Utilization** |  |  |  |
| IABP Only | 87 (30.9%) | 97 (34.4%) | 0.94 |
| Impella devices Only | 30 (10.6%) | 31 (11.0%) |  |
| Medical Therapy Only | 165 (58.5%) | 154 (54.6%) |  |
| **Major Bleeding** | 44 (15.6%) | 40 (14.2%) | 0.72 |
| **Mortality** | 88 (31.2%) | 84 (29.8%) | 0.78 |

Among these two clinical subgroups of n=282 patients each, MCS utilization, bleeding, and mortality rates were not significantly different. Patients with a benefit with IABP had a significantly higher rate of emergent PCI (87.2% vs 39.0%, p < 0.001) and a significantly higher rate of STEMI present on initial ECG (85.1% vs 50.0%, p < 0.001). Patients with a benefit with Impella use were on average younger (66.6 ± 14.9 years vs 70.7 ± 10.0 years, p < 0.001), presented with higher systolic blood pressure (130.6 ± 58.4 mmHg vs 102.0 ± 52.5 mmHg, p < 0.001), higher rate of salvage PCI (12.4% vs. 6.0%, p < 0.001), higher initial creatinine (2.41 vs. 1.25 mg/dL, p < 0.001) and more likely to present with NSTEMI This population tended to have a higher degree of proximal LAD stenosis (52.1% vs 26.3%, p < 0.001). Finally, this population had significantly higher incidence of NSTEMI (48.6% vs. 13.1%, p < 0.001).

### Interpretation of Factors Associated with Risk

While the risk differences provide a prospective view, model interpretation was performed on the retrospective models with SHapley Additive exPlanations (SHAP) values and plots in order to understand risk factors and model performance^20, 21^ and this analysis is explained in the **Supplementary Material**. In brief, age was the most strongly informative feature, as ranked by SHAP, followed by initial creatinine and history of cardiac arrest. Higher age, creatinine, and history of cardiac arrest were all associated with increased risk. SHAP contributions for the model incorporating MCS device utilization found that MCS device utilization was the fourth most informative feature.

## Discussion

While prior findings (and this work) noted that overall in the population, Impella devices were associated with higher mortality risk compared to IABP^1, 10–12^, some patients with certain clinical profiles were, in fact, associated with lower risk and different variables had different impact on this risk, illustrating significant patient heterogeneity. Specifically, our analysis identified 282 patients with a decreased risk of mortality when hypothetically managed with Impella devices as compared with IABP. Those patients were on average younger, presented with higher systolic blood pressure, higher rate of salvage percutaneous coronary intervention, higher initial creatinine, and lower hemoglobin. While Impella devices were associated with higher mortality risk overall, certain clinical profiles were associated with lower risk, illustrating patient heterogeneity.

These findings align with recent trials identifying subpopulations where MCS device utilization led to improved outcomes ^13^. This analysis suggests that patients with certain clinical features, or a combination of factors, may have a larger benefit or a harm with different MCS strategies, highlighting the need for personalized estimation of risk and benefit to support tailored decision-making. Once the XGBoost model has been trained, it is able to produce both an estimate risk of mortality and an estimation for how that risk would change given different MCS treatment choices. SHAP can display the factors that lead to these individual risks, and an implementation tool could allow for model transparency to enable researchers and clinicians to understand what nonlinear interactions of factors drive the overall estimated risk.

It is interesting to note that when comparing the two populations (which represent those with potential benefit from Impella devices compared to IABP and an equal number with the highest risk from Impella devices compared to IABP) in **Table 5**, no significant difference is seen with outcomes relative to MCS utilization. This implies that the factors that the model uses to estimate mortality may not be fully understood in clinical practice. If a clear difference in utilization between these groups were present, it would indicate that the model’s training primarily focused on clinician treatment patterns. However, since no difference in MCS utilization can be seen between these groups, it is more plausible that the model is instead identifying a previously unrecognized interaction between patient factors. Decision tree methods are capable of exploring subpopulations and identifying similar patients based upon measured factors. While limited, the completeness of the registry data allows for comprehensive exploration, similar to work that explores subpopulations in more general clinical settings ^22^.

It is possible this is a reflection of the challenges clinicians face integrating multiple features in real-time with an unstable patient, as well as a need for additional risk factors measured for understanding the association with treatment and risk (discussed in limitations). For the former, a further investigation is needed to test the findings of these models and these hypotheses on patients associated with lowered risk, to refine understanding of the relationship between MCS utilization and mortality in AMI-CS patients in real-time while treatment decisions are made.

### Limitations

This work has several limitations. First, this is a retrospective analysis that is subject to unmeasured confounding. However, the tails of the specific distribution in **Figure 2** may help inform randomized control trials to determine if and which patients benefit from IABP, Impella devices, or no MCS device. We could not distinguish specific models of MCS devices used (such as Impella device models) nor did we consider groups that involved combinations of various MCS devices. While the sites selected were those that prescribed both IABP and Impella, it did not adjust for such clinical sites that strongly preferred one to the other. Therefore, there is a potential for selection bias in the findings, given the limited focus on those that generally prescribed both MCS devices, not including any locations that may have prescribed more or multiple MCS devices not considered in this analysis. Finally, the SHAP feature importance plots show individual feature rankings and associations with outcomes but do not clearly illustrate the higher order interaction between the terms. While figures were provided that visualize two-term interactions, trees of depth 6 allow for potentially 6 terms to interact together to better estimate risk of mortality. This association, however, is challenging to identify and illustrate.

### Conclusion

Advanced machine learning models, which can classify patients into subgroups of risk as driven by particular factors, better model risk of mortality in heterogeneous patient populations. These risk factors, and their associated importance in mortality prediction when interacting with MCS device utilization, identified cohorts of individuals where selection of particular MCS utilization for treatment may be associated with improved risk of mortality. This information could help guide potential cohort selection for future randomized control trial design in order to better identify best practices for treatment decisions in the heterogenous group of patients with AMI-CS.

## Online Methods

### Data Sources

Our data sources were linked American College of Cardiology’s National Cardiovascular Data Registry CathPCI and Chest Pain-MI registries, as previously described^1, 7^. The linkage provided full episode of care detail because of complementary data from the CathPCI registry and detailed MCS utilization data from the Chest Pain-MI registry. These registries have high data quality standards, including requiring data completeness^23^. From the matched cohort, we eliminated episodes of care from any hospital without at least one of each of Impella device or IABP within the AMI-CS cohort, to ensure cases were at sites with capability to use any of these therapies. Additionally, we eliminated episodes of care where a given patient received an MCS other than Impella devices or IABP or where a given patient received multiple MCS devices.

### Outcomes

The primary outcome was in-hospital mortality. A secondary outcome was in-hospital major bleeding. Major bleeding was defined as in prior work^1, 7^ as a decline in hemoglobin of at least 3 g/dL; transfusion of whole blood or packed red blood cells; procedural intervention/surgery at bleeding site to treat the bleeding; or documented or suspected retroperitoneal bleed, gastrointestinal bleed, genitourinary bleed, or a bleed in a location not specified elsewhere^24^.

### Statistical Analysis

We trained machine learning models to estimate risk of in-hospital mortality using the extreme gradient boosting machine (XGBoost)^14^. XGBoost is an efficient implementation of the gradient boosting machine (GBM) algorithm^25^ with several algorithmic and hardware enhancements leading to better scalability, speed, and performance accuracy. An important enhancement in XGBoost is the implementation of the least absolute shrinkage and selection operator (LASSO) and the ridge (Ridge) regularization techniques^26^, which can penalize large and complex models to prevent overfitting. The XGBoost model has been shown to improve model performance in the NCDR registries^15, 16^. We used a nested approach for model development, hyperparameter tuning, and evaluation. In the primary evaluation (for model performance), we conducted a stratified, five-fold cross-validation, with each fold splitting the data 80% for model training and development and 20% for model testing and analysis. For that 80% selected for training, we conducted an inner evaluation for hyperparameter tuning, through a stratified twenty-fold cross-validation. Here, 95% of the data were used to train and 5% to test to select optimal model hyperparameters. The hyperparameters of our model were the learning rate (searched from 0.1 to 0.3), the maximum depth of each tree (1,2,3,6), and the number of trees used in the final model (50, 100, 500, 1000). Other parameters were left at their default values. Further details are provided in the **Supplementary Material**. Model performance was assessed using the area under the receiver operating characteristic curve (AUROC), with a 95% confidence interval.

### Covariates and Feature Selection

To develop a parsimonious and interpretable risk model, we followed a stepwise process which included both clinically driven and data-driven steps for feature elimination. We began with a large set of variables used for propensity matching in prior research^1^. These included patient demographics, medical history, clinical presentation, laboratory values, administered medications, procedural characteristics, and coronary anatomic data. Binary variables with missing data were coded as “no”, while missing categorical variable levels were coded as “no” or “other”, and continuous variables imputed by the mean, though rates of missing data were extremely low^23^.

We then began our process of eliminating variables. We first eliminated left ventricular ejection fraction because this measure is not always available at the time of treatment decision. We eliminated peri-procedural medication utilization to avoid biasing the model based on treatment decisions. We then methodically eliminated features using backwards selection: we trained a model on all features, and then eliminated the least informative variables as ranked by XGBoost’s internal importance ranking. We continued this removal process until removal of the least-important feature resulted in an AUROC below the 95% confidence interval surrounding the original model’s AUROC. This was done in order to both minimize model overfitting and to allow improved generalization with a more parsimonious model.

### Staging of Models

We then built additional models that considered the selection of MCS utilization for treatment. While the first model used only pre-treatment decision factors, the second model added MCS utilization, either Impella device or IABP. This model was used to interpret the association of MCS utilization with both mortality and changes in feature importance of variables. This design allows for the comparison of associations of each covariate with in-hospital mortality both with and without knowledge of MCS device utilization.

### Estimated Differences in Risk of Mortality

We next used a simulated prospective approach to estimate risk changes with different MCS treatment choices. This was accomplished using the second-tier model (Baseline + MCS Utilization) and comparing differences in estimated risk for all test patients. The change in the estimated risk reflects how the treatment effect of MCS utilization is heterogeneous or varies across individuals and their characteristics. For each individual patient, a set of risk estimates (the probability of an outcome given MCS utilization and covariates) was generated. This set of estimates was generated by imputing MCS utilization to each of its two possible values, independent of the intervention the patient actually received. Two risk estimates are therefore generated for each subject: 1) an estimate for if that subject received Impella device only, and 2) an estimate if that same subject received IABP only illustrated in **Figure 1**. The risk difference for two interventions was estimated by subtracting these values. The risk difference for Impella device vs IABP was estimated as:

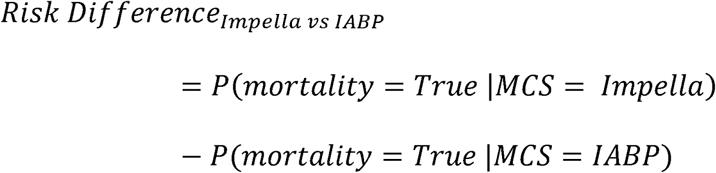

**Figure 1.**
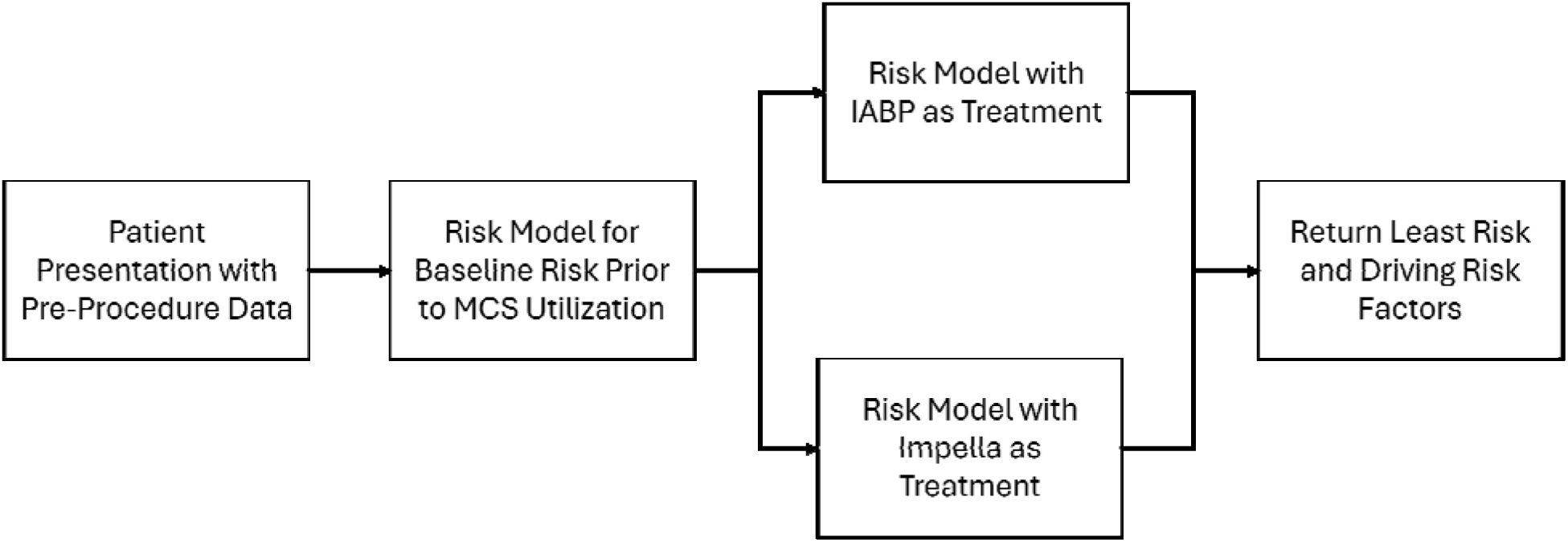
Flowchart of Risk Models and Treatment Decision Making. This figure illustrates the model development process and how a final risk score is calculated.

**Figure 2.**
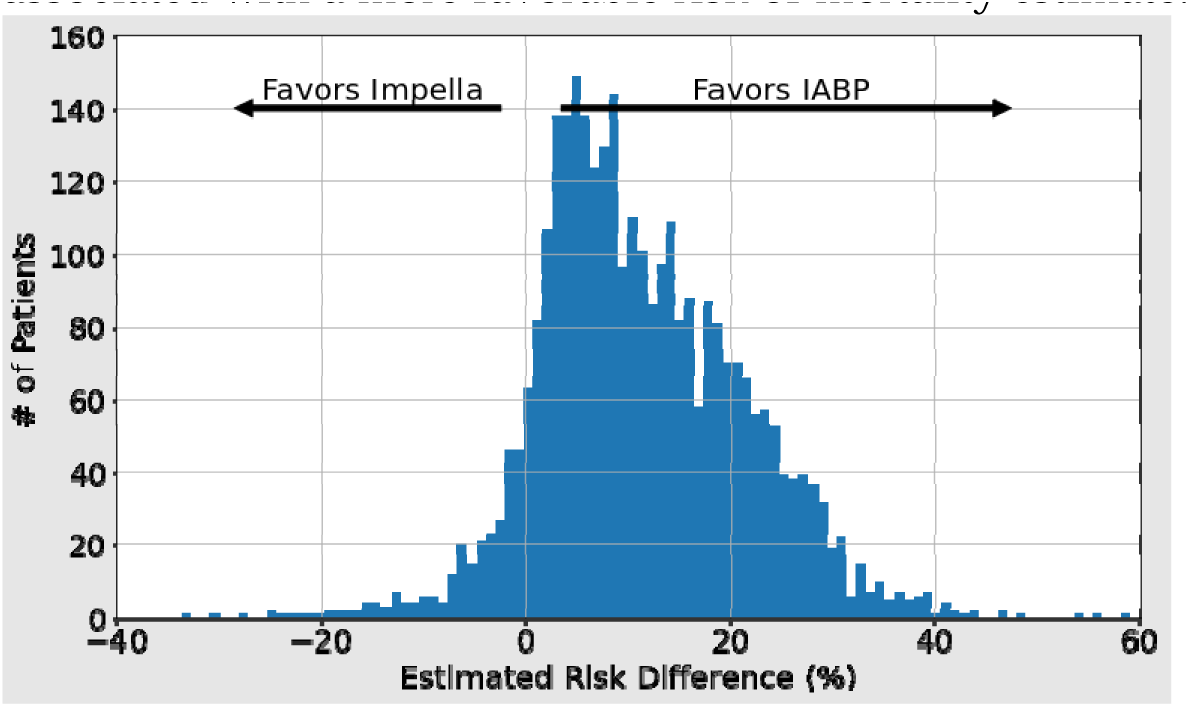
Estimated Mortality Risk Difference of Using IABP vs. Impella Devices. This figure illustrates the subset of patients in which the use of either IABP or Impella devices is associated with a more favorable risk of mortality estimate.

Here a positive estimated risk difference indicates a given patient’s features are associated with higher risk of mortality if treated with Impella device rather than IABP. The distribution of estimated risk differences was visualized using a histogram. To compare the strongest changes in predicted outcome, the tails of this distribution were compared. Among the populations of these tails, patient characteristics were compared with Mann-Whitney U tests for continuous variables and Fisher’s Exact Test for categorical variables.

### Interpretation of Risk Factors

While the risk differences provide a prospective view, model interpretation was performed on the retrospective models with SHapley Additive exPlanations (SHAP) values and plots in order to understand risk factors and model performance^20, 21^ and this analysis is explained in the **Supplementary Material**.

All data analyses was performed in Python 3.6 and R4.1, with the packages XGBoost 1.3.3^14^, SHAP 0.38.1^27^, and scikit learn (sklearn) 0.24.1^28^.

## Supporting information

Supplemental Methods and Results

## Data Availability

All data from this study are available online upon reasonable request to the American College of Cardiology at https://cvquality.acc.org/NCDR-Home

## Acknowledgements

### Funding/Support

This work was supported by the Food and Drug Administration (FDA) of the U.S. Department of Health and Human Services (HHS) as part of a financial assistance award [FD005938] totaling $92,921 with 100 percent funded by FDA/HHS. The contents are those of the authors and do not necessarily represent the official views of, nor an endorsement by, FDA/HHS, or the U.S. Government.

### Potential Competing Interests

Dr. Mortazavi currently receives research support through the National Science Foundation (IIS 2014475), and the National Institutes of Health (1 R01 HL167858-01A1, 1 R01 DK136414, and 1 R01 HL151240-01). Dr. Mortazavi is an expert witness at the Kirkland and Ellis Law Firm for matters unrelated to this manuscript. Dr. Mortazavi was an expert witness at the request of Abbott’s attorneys, the McAndrews Held and Malloy Law Firm, in a case that was settled in December of 2024. He currently serves as an Associate Editor for the Journal of the American College of Cardiology.

Dr. Ross currently receives research support through Yale University from Johnson and Johnson to develop methods of clinical trial data sharing, from the Food and Drug Administration for the Yale-Mayo Clinic Center for Excellence in Regulatory Science and Innovation (CERSI) program (U01FD005938), from the Agency for Healthcare Research and Quality (R01HS022882), and from Arnold Ventures; formerly received research support from the Medical Device Innovation Consortium as part of the National Evaluation System for Health Technology (NEST) and from the National Heart, Lung and Blood Institute of the National Institutes of Health (NIH) (R01HS025164, R01HL144644); and in addition, Dr. Ross was an expert witness at the request of Relator’s attorneys, the Greene Law Firm, in a qui tam suit alleging violations of the False Claims Act and Anti-Kickback Statute against Biogen Inc. that was settled September 2022.

Dr. Ngufor reports that, in the past 36 months, he has received research support through the Department of Defence (DOD) Prostate Cancer Research Program (PCRP) Health Equity Research and Outcomes Improvement Consortium (HEROICA) Award W81XWH2210968. No other conflicts of interest exist.

Dr. Curtis has a contract with the American College of Cardiology for his role as Chief Scientific Advisor of the American College of Cardiology’s National Cardiovascular Data Registry. He holds equity interest in Medtronic.

Dr. Krumholz, in the past three years, has received options for Element Science and Identifeye and payments from F-Prime for advisory roles. He was a co-founder of and held equity in Hugo Health. He is a co-founder of and holds equity in Refactor Health and ENSIGHT-AI. He is associated with research contracts through Yale University from Janssen, Kenvue, Novartis, and Pfizer. He is the Editor-in-Chief of the Journal of the American College of Cardiology.

## Footnotes

1 The models performed similarly across all folds as demonstrated by the confidence intervals on the AUROCs, so we present a single fold, without loss of generality. The additional folds are illustrated in the **Supplementary Material.**

## Notes

### Author Declarations

Ethics committee of Yale University waive ethical approval of this work under IRB #0607001639.

