## Supplemental Methods and Results for "Modeling In-Hospital Mortality Among Patients Undergoing Percutaneous Coronary Intervention with Acute Myocardial Infarction Complicated by Cardiogenic Shock Receiving Mechanical Circulatory Support"

*Hyperparameter Tuning for XGBoost Trees*

In order to develop the optimal model for risk estimation, many machine learning models have hyperparameters that must be tuned; that is, for each model we must select the parameters and structure of the model to perform best. For XGBoost, the primary parameters that we searched were the depth of each tree estimator (in other words, how many potential levels of interaction between variables were allowed), the number of estimators (trees) in the final ensemble voting model, and the learning rate, the rate at which the algorithm decides additional trees are needed and samples re-weighed in training.

For this work, we considered the maximum depth of the tree to range from 1 (direct, linear interaction between terms and final estimation), 2, 3, and default value of 6. This does not mean each tree must have a depth of 6, but that 6 levels of interactions are allowed if this helps partition the space and improve risk estimates in training. We also considered a wide array of a number of estimators, from 50 to 1000 trees to use in the final weighted ensemble prediction. The model, when using 1000 trees, has a weight associated with how much each tree contributes to the final risk estimate. Finally, we also adjusted the learning rate from 0.1 to 0.3. These models work best when learning slowly (low learning rate), only trusting that they have produced a risk estimate for a sample in the training set well if multiple trees correctly estimate the outcome for that sample.

Finally, to avoid data leakage in the final model, we identified the optimal hyperparameters from only the training set. In the initial five-fold cross validation – 20% of the data is held out for testing. For the remaining 80% we simulate training and testing the model, taking this data and further partitioning it. This allows us to score how well models with different selected hyperparameters perform, to best estimate how they will perform on unseen data (the held-out test set). We repeated this 20 times, selecting the best hyperparameters across these tests, and finally training a complete model with these fixed hyperparameters (depth 6, 100 trees, with a learning rate of 0.3).

*Interpretation of Risk Factors*

The SHAP values provide a model-agnostic approach to interpreting the strength of association of each variable with mortality, including an estimation of ranked feature importance across all participants and values of the specific features. Specifically, the SHAP method assigns each variable in the model an importance score that measures the contribution of the variable towards individual risk predictions. A positive SHAP value for a variable indicates it contributes to greater event likelihood, while negative value indicates a contribution to lower event likelihood. Given that the underlying XGBoost model is nonlinear, the SHAP values provide an interpretable explanation of nonlinear effects.

*SHAP Contributions for Additional Folds*

The SHAP contributions in the primary manuscript represented the first fold. We illustrate the other four folds here to demonstrate the stability of the model in cross-validation and ability to generalize the findings of one fold as an appropriate representation of hold-out performance. **Supplementary Figures 2-5** illustrate the additional folds (2-5) of base model and **Supplementary Figures 6-9** illustrate the additional folds after including MCS utilization (folds 2-5). Note that at most only a few variables swap positions across folds with each other, usually limited to a move in importance order by one, demonstrating that they’re equally important.

*Explanatory Plots*

SHAP contributions of the base model are shown in **Figure 3**. Age was the most strongly informative feature, as ranked by SHAP, followed by initial creatinine and history of cardiac arrest. Higher age, creatinine, and history of cardiac arrest were all associated with increased risk.

SHAP contributions for the model incorporating MCS device utilization found that MCS device utilization was the fourth most informative feature (**Figure 4**). However, as shown by the coloring (red: no MCS device, blue: IABP, and purple: Impella), the average trend does not apply to every patient, identifying the existence of heterogeneity in the patient population. To better visualize the impact of the three possible values of MCS device utilization, **Figure 5** shows the same information about MCS as a single-variable dependence plot for the average trend^22^, while **Figures 6 and 7** illustrate the relationship in terms of interactions. While Impella is associated with higher risk of mortality on average (**Figures 4 and 5**), it is associated with lower risk in some patients with cardiac arrest **(Figure 6)** and mixed with patients with multivessel disease **(Figure 7)**.

**Supplementary Figure 2. SHAP contributions of each base model feature in predicting in-hospital mortality in Fold 2.**

Red color indicates the higher value for the feature, blue the lower. The further right is associated with higher risk of mortality, the further left is the lower risk of mortality. Binary variables of no/yes (0/1) are coded as blue/red.

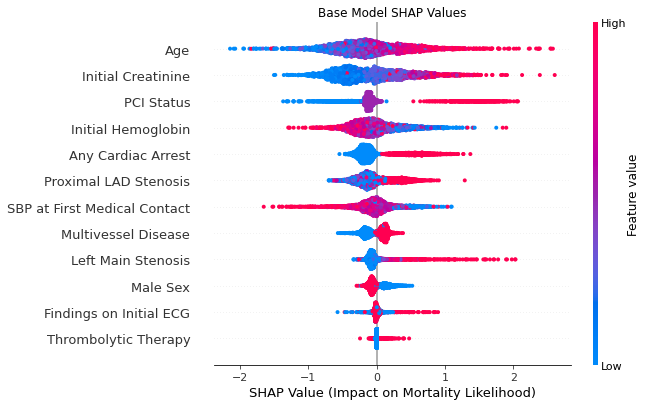

**Supplementary Figure 3. SHAP contributions of each base model feature in predicting in-hospital mortality in Fold 3.**

Red color indicates the higher value for the feature, blue the lower. The further right is associated with higher risk of mortality, the further left is the lower risk of mortality. Binary variables of no/yes (0/1) are coded as blue/red.

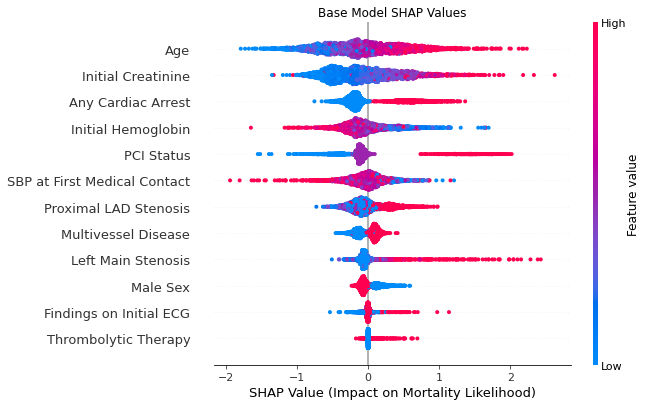

**Supplementary Figure 4. SHAP contributions of each base model feature in predicting in-hospital mortality in Fold 4.**

Red color indicates the higher value for the feature, blue the lower. The further right is associated with higher risk of mortality, the further left is the lower risk of mortality. Binary variables of no/yes (0/1) are coded as blue/red.

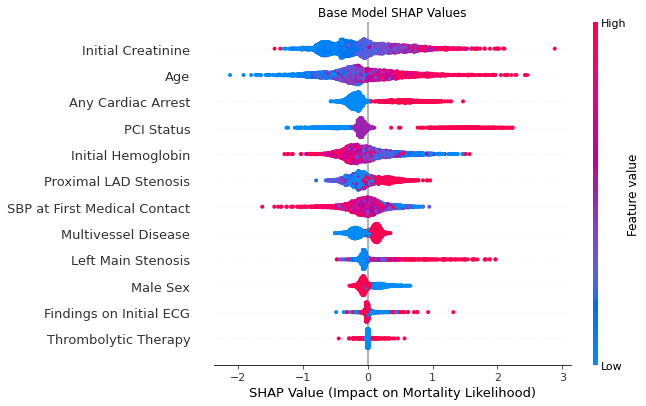

**Supplementary Figure 5. SHAP contributions of each base model feature in predicting in-hospital mortality in Fold 5.**

Red color indicates the higher value for the feature, blue the lower. The further right is associated with higher risk of mortality, the further left is the lower risk of mortality. Binary variables of no/yes (0/1) are coded as blue/red.

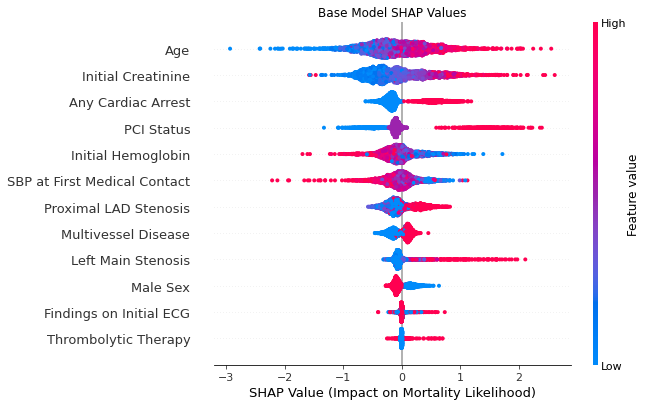

**Supplementary Figure 6 SHAP contributions incorporating MCS device utilization for Fold 2**. Notice the inclusion of Device Utilization (2^nd^ feature), coded so that red is no MCS device, blue is IABP, light purple is Impella, and dark purple is Multiple/Other.

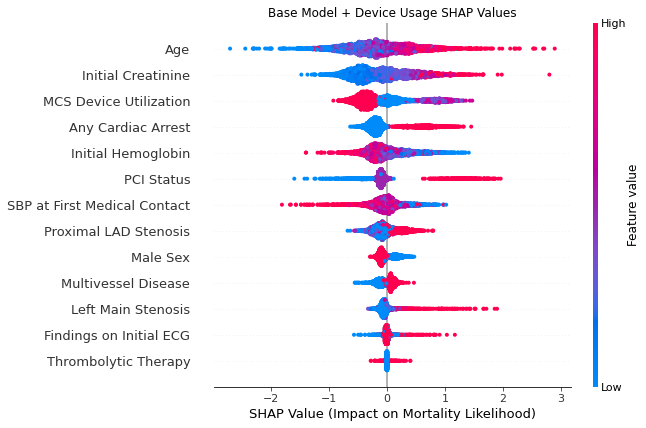

**Supplementary Figure 7 SHAP contributions incorporating MCS device utilization for Fold 3**. Notice the inclusion of Device Utilization (2^nd^ feature), coded so that red is no MCS device, blue is IABP, light purple is Impella, and dark purple is Multiple/Other.

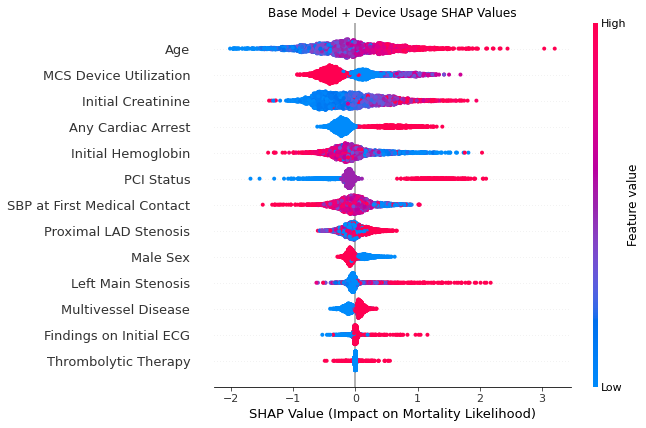

**Supplementary Figure 8 SHAP contributions incorporating MCS device utilization for Fold 4**. Notice the inclusion of Device Utilization (2^nd^ feature), coded so that red is no MCS device, blue is IABP, light purple is Impella, and dark purple is Multiple/Other.

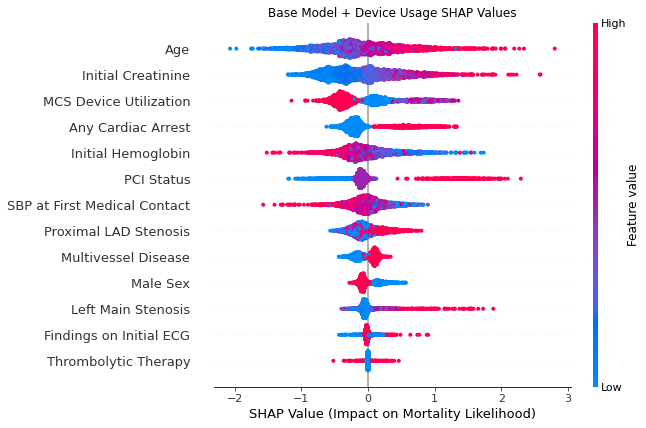

**Supplementary Figure 9 SHAP contributions incorporating MCS device utilization for Fold 5**. Notice the inclusion of Device Utilization (2^nd^ feature), coded so that red is no MCS device, blue is IABP, light purple is Impella, and dark purple is Multiple/Other.

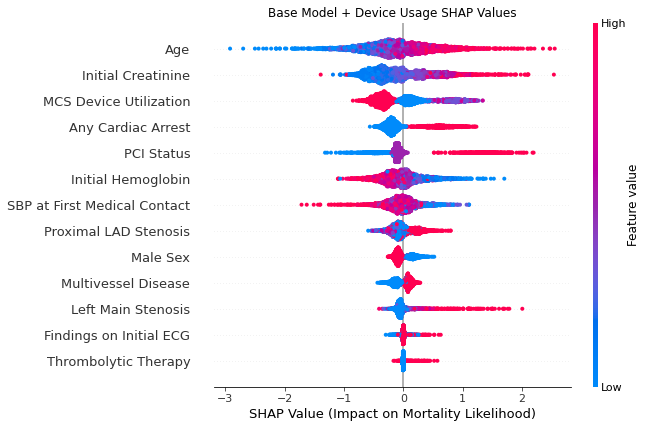

**Figure 3 SHAP contributions of each base model feature in predicting in-hospital mortality.**

Red color indicates the higher value for the feature, blue the lower. The further right is associated with higher risk of mortality, the further left is the lower risk of mortality. Binary variables of no/yes (0/1) are coded as blue/red. The y-axis lists the variables in order of importance from top to bottom.

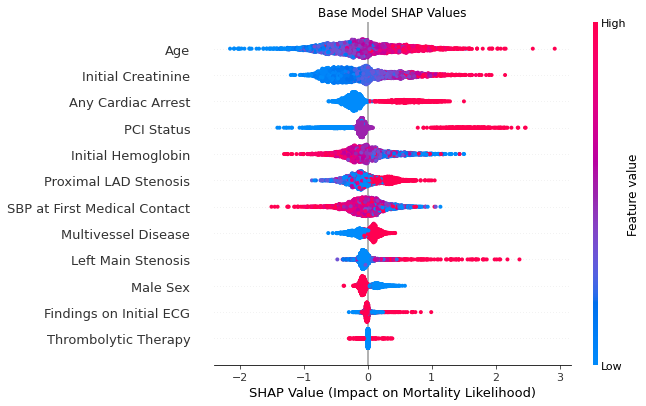

**Figure 4 SHAP contributions incorporating MCS device utilization**. Notice the inclusion of Device Utilization (2^nd^ feature), coded so that red is no MCS device, blue is IABP, and purple is Impella.

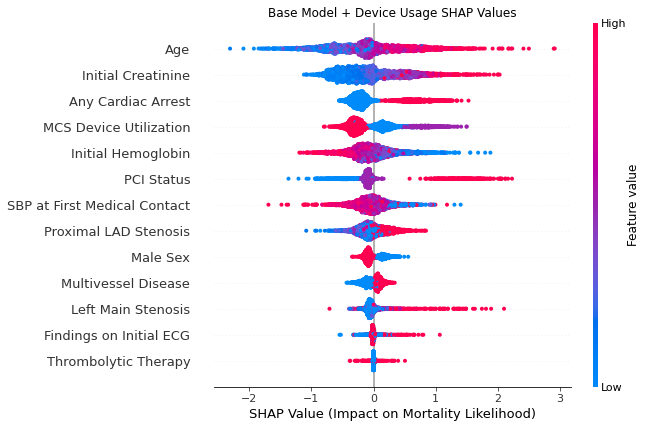

**Figure 5 SHAP effect contributions as a function of MCS device utilization.** This plot shows the data from the “MCS Device Utilization” row in Figure 3, which indicates, on the average, IABP is associated with a higher risk of mortality, Impella is more strongly associated with a higher risk of mortality, and medical therapy alone is associated with a lower risk of mortality.

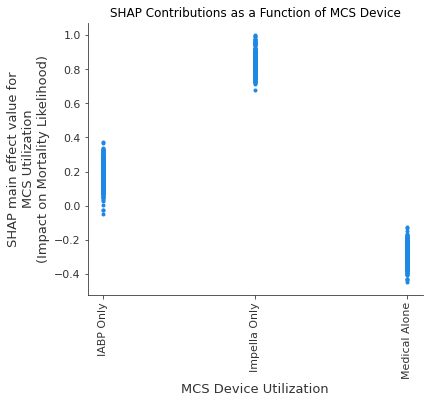

**Figure 6 – Interaction comparison for Cardiac Arrest and MCS Utilization**

This figure illustrates the interaction and risk estimate between use of MCS device and whether the patient had any cardiac arrest. The figure would suggest IABP or Impella are associated with better outcomes in any case where the patient had cardiac arrest, but that medical therapy alone is associated with worse outcomes in any case with cardiac arrest.

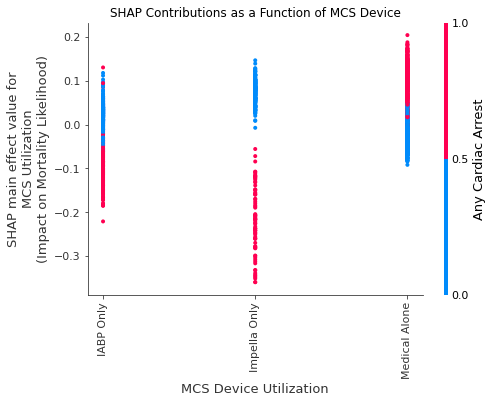

**Figure 7 – Interaction between MCS Utilization and Multivessel Disease.**

This figure illustrates the interaction and risk estimate between MCS device utilization and multivessel disease. This comparison suggests a strong association between better outcomes and IABP in cases with multivessel disease, as opposed to Impella.

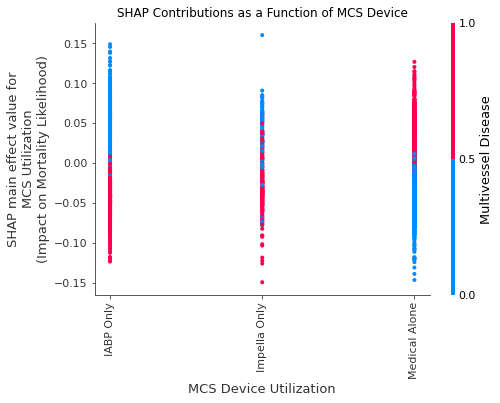
